# Assessing community factors associated with Antiretroviral Therapy (ART) defaulting among youth accessing HIV care in Mzimba District, Malawi

**DOI:** 10.1101/2023.08.11.23293981

**Authors:** Isaac Paul Kasalu, Mathews Lazaro, Idesi Chilinda

**Affiliations:** Mzimba District Hospital, P.O. Box 131, Mzimba, Malawi; Faculty of Applied Health Sciences, Kamuzu University of Health Sciences, P/Bag 1, Lilongwe, Malawi; School of Nursing, Community Health Nursing Department, Kamuzu University of Health Sciences, P/Bag 1, Lilongwe, Malawi

**Keywords:** Antiretroviral therapy, Defaulting, community social support, youth

## Abstract

**Introduction/Background:** ART defaulting has been associated with increased morbidity and mortality of HIV positive youth. Youth that default tend to develop drug resistance and resurgence of opportunistic infections. They can also transmit drug resistant strains of HIV to others through unprotected sexual intercourse. This study, therefore, aims at assessing community factors influencing ART defaulting amongst HIV positive youth enrolled on ART in Mzimba District.

**Material and Methods:** This study utilized a quantitative, case control design. A sample of 411 HIV positive youth (n=137 cases and n=274 controls) attending an HIV care clinic was recruited. Both ART defaulters and non-defaulters were enrolled using random sampling technique. Data were collected using a structured questionnaire and analyzed using the Statistical Package for Social Scientists (SPSS) version 20.0. Descriptive statistics were used to provide count, frequencies, proportions and ranges while inferential statistics were used to establish association between dependent variable with independent variables.

**Results:** Results from this study indicate that lack of community social support (*p*< 0.001, OR: 11.257, 95% CL: 6.782-18.686), long distance to ART clinics (*p*: 0.002, OR: 2.454, 95% CL: 1.511-3.985) and migration of the youth other countries (*p:*0.001, OR: 35.661, 95% CL: 4.675-272.049) are statistically significant factors to ART defaulting among the study participants who are youth in Mzimba District in Malawi.

**Conclusion:** The study conclusively proved that lack of community social support, forgetfulness and going to RSA for employment influenced defaulting to antiretroviral therapy among the youth in Mzimba, Malawi. The researcher recommends introduction of community ART outreach programs to help in reducing defaulting among the youth. Further, policy review to provide for technology that enables free ARVs regardless of national identity (ID) documents requirements and cross-border collaboration in managing HIV among the youth on ART between countries to ensure continuity of care.

## Introduction/Background

Antiretroviral therapy (ART) defaulting remains a major challenge for youth living with Human Immunodeficiency Syndrome (HIV) globally and in Malawi. An average of 38% ART defaulting rates have been reported among the youth globally ^1^. In Malawi, ART defaulting has been reported at 15.5% ^2^. Although Malawi has made positive progress in reducing ART defaulting rate across key populations^3^, this has not been the case with the youth^4, 5^. This paper considers the youth as ages from 15 to 35 years as defined by the Malawi National Youth Policy. ART defaulting makes an individual susceptible to opportunistic infections, increases both the risk of drug resistance, HIV transmission and mortality^5^.

Lack of community social support in ART has been linked to increasing treatment defaulting among clients with HIV in many countries ^6, 7^. Defaulting carries diverse meanings among different authors. Others define defaulting as a situation where a client enrolled on ART missed his/her last clinic appointment by more than 1 month ^8^. This study defines defaulting according to the Malawi HIV Guidelines, as an HIV positive individual who fails to report for new drug supplies at 2 months when he or she is expected to have run out of ARVs^9^. This practice deprives the individual of drugs to take on next scheduled times. ART defaulting contributes to early disease progression and increased mortality related to HIV infections ^10^.

Reducing ART defaulting is a priority for Malawi government towards achieving the 95-95-95 targets. The fast track 95-95-95 targets were set aside by the United Nations general assembly in 2016 with the aim of eliminating the burden of HIV in the world^11^. These targets are intended to make sure that 95% of HIV positive individuals know their sero-status, 95% of those receive ART, and 95% of those enrolled on ART achieve viral suppression. However, evidence has shown that achieving viral suppression is of great concern due to ART defaulting among the youth. This makes the dream of achieving the last 95% as a challenge. Addressing challenges associated with ART defaulting among the youth may not only help the nation to realize its goals in the set target but also assist to reduce the mortality due to HIV related illnesses.

Literature from several sources in the world indicate that the youth face a lot of challenges that affect their participation in ART programs^7, 12, 13^.These could be social, physical, psychological and health system related in nature. Some studies conducted in Malawi cited examples of the social factors among the adults in Malawias lack of community social support and fear of disclosure of HIV status^8, 14^. Some of the physical factors cited include experiencing side effects of ART while health systems factors include poor reception by the care provider, and being concerned with privacy among others^15–17^.Nonetheless, there is a gap in identifying specific factors contributing to ART defaulting among the youth in Mzimba District.

Evidence has shown that some youth in Mzimba District default ART treatment due to long distances to the heath facility whilst others migrate to the Republic of South Africa (RSA) in search for greener pastures among other reasons. Being in a foreign land, some youth have limited access to medical care including ART supplies^18^. While some of the relatives are able to collect ART drugs and send to RSA, others do not have such support. Eventually, these clients may default treatment. Nevertheless, no research study has been conducted to assess the effects of these factors on ART program in Mzimba District. Therefore, this study seeks to assess community social factors contributing to ART defaulting in Mzimba District. A better understanding of the community social factors associated with ART defaulting would provide a basis for planning evidence based interventions for communities in Mzimba District to support the youth on treatment.

## Material and Methods

### Research Design

A case–control study design was utilized in this study. Cases were all youth that defaulted ART while controls were all youth that remained on treatment. This research design compared the occurrence of ART defaulting in cases and controls to determine if there was any association. The researchers intended to determine if community social factors were associated with ART defaulting among the youth by comparing occurrence of these factors and client’s failure to return to the clinic on the next appointment date as an outcome.

### Study time and population

The study was conducted from March 2020 to February 2021. The population targeted in this study comprised of all HIV positive youth enrolled on ART treatment in Mzimba District.

### Inclusion and exclusion criteria

All HIV-positive youth aged 15–35 years who were enrolled in the ART program from July 2018 to June 2020 were eligible for inclusion in the study. This study included both defaulters and non-defaulters who provided informed consent. For those below the age of 18 years, consent was sought from their parents or guardians, and then an assent was signed to participate in the study. Excluded in this study were all HIV-positive youth diagnosed and recruited in ART programs before July 2018 or after June 2022. Also excluded were all HIV-positive youth on ART who were transferred out of the health facility. Furthermore, all HIV-positive youths who did not meet the age criteria of 15–35 years and all non-HIV-positive youths attending other health facility services were not included in this study.

### Sampling, sample size and sampling technique

The study required 411 participants in the ratio of 1: 2 for cases and controls respectively. Therefore 137 cases and 274 controls were recruited. The researcher extracted client details from ART register, ART registration cards and ART computers to locate them and verify their eligibility for the study. Extracted details included: physical address and phone numbers, last visit date to the clinic and pharmacy, next scheduled return date for the clinic and pharmacy appointment, deaths, transfer-in date and transfer-out. Cases and controls were matched on the perspective of health facility from where the youth accessed ARVs and time under study. A two-stage cluster sampling approach was used to select the participants in phases. The first phase that determined health facilities for the study used simple random sampling. Names of the health facilities were written on a piece of paper, folded and placed in a closed box. A volunteer was requested to draw one paper at a time, after shaking the box. The drawn papers were read and listed on an A4 piece of paper. The process was repeated until four facilities were determined in a similar manner. These were: Mzimba District Hospital, Embangweni Hospital, Jenda and Euthini health centres. Phase two used stratified random sampling to recruit participants (cases and controls) from the identified health facilities in the ratio of 1:2. Four cartons were made available at each of the identified facilities containing identity numbers of clients. To determine cases, a volunteer randomly drew identity numbers from the carton, one at a time after shaking. The process was repeated for the non-defaulting youth till the specific sample size per health facility was achieved.

### Data collection methods, tools and data management

The researcher used medical records such as ART registers and facility computers to identify individual study participants. Dates in which data were accessed differed by facility. Table 1.0 provides the exact dates when these records were accessed.

**Table 1.0:** Dates for reviewing medical records for research purposes.

| Health Facility | Data access dates |
| --- | --- |
| Euthini | 24-08-2020 |
| Embangweni | 25-08- 2020 |
| Jenda | 26-08-2020 |
| Mzimba | 27, 28- 08- 2020 |

A structured questionnaire with close-ended questions was used to collect data from eligible participants. Data collection was done from 1^st^-30^th^ September 2020. Socio-demographic data which included participant’s age, sex, occupation, marital status and level of education were collected (Table 1.0). More information was sought on stigma and discrimination, distance to health facility and cost of travel to the clinic. The questionnaire also included reasons that led to client’s failure to attend clinic as scheduled. Five data collectors (including the lead author and four qualified nurses) were used during this study. A three-day training was provided to the four nurses to familiarize them to the study methodology and measurement of defaulting and its associated factors. All data collected were stored in a lockable filing cabinet placed in the maternal and child health (MCH) Office. Keys of this cabinet were only accessed by the researcher. Data in computer were protected by the passwords known only to the researcher.

### Ethical and Cultural Considerations

Approval to conduct the study was sought from the College of medicine research and ethics committee (COMREC). Further permission was pursued from the Director of Health and Social Services (DHSS) of Mzimba South and the Medical Director of Embangweni Mission Hospital for the study to be conducted in their health facilities. In addition, a written informed consent was obtained from the eligible participants. Parents or guardians provided a written consent for their eligible minor participants below the age of 18. A written assent was also sought from the respective youth before the interview. Participants or guardians were given information sheet for the study to read and they were requested to sign if they understood and agreed to participate on their own free will. The participants were assured that involvement in the study was voluntary and that they were at liberty to participate in the study or not without giving reasons for their choice. Further, they were informed that they were at liberty to drop out of the study at any point during interview without giving reasons.

Data collection took place in private rooms with closed doors to ensure both visual and audio privacy and confidentiality. Codes were used to identify clients on the questionnaire to ensure anonymity. Participants were guaranteed that their information would be kept with strict confidence throughout the study. The researcher treated with confidentiality all the information given by the respondents during the interviews.

### Data Analysis

Data were entered on statistical package for social scientists (SPSS) software version 20.0 for cleaning and analysis. The dependent variable was ART defaulting. Two levels of dependent variables were analyzed, namely: defaulting and not defaulting. Both descriptive and inferential statistics were used. In descriptive analysis, data analyzed were organized and presented in form of frequency distribution tables. For inferential analysis, Chi-square testing was performed by cross tabulating dependent variable with independent variables. Factors which were statistically significant were included in binary logistical regression to assess their influence on ART defaulting. A 5% level of significance was used in all statistical tests in this study

## Results

As seen in table 2.0, a total of 411 participants were interviewed from 4 health facilities namely: Mzimba District Hospital, Embangweni Mission Hospital, Jenda and Euthini Health Centers. The majority of participants 25.5% (n=105) were in the age range of 31-35 yeas. This group comprised of 6.8% (n=28) cases and 18.7% (n=77) controls. The least age group to participate in the study was 21-24 years 13.9% (n=57). About 29% (n=119) of the participants were males whilst 71% (n=292) were females. Most of these youth 59.4% (n=244) attained primary school education and the majority of them 45% (n=185) were married. Many youth 90% (n=369) were Christians. In terms of means of travel to the health facility: the majority of the participants 72.5% (n=298) would walk to the health facility to access health services and 43.3% (n=178) of them took less than 30 minutes to get to the health facility.

**Table 2.0.** presents the socio-demographic and economic characteristics of the study participants.

| <u>Variable</u> | <u>Frequency/Percentage</u> | <u>Cases</u><br><u>(n=411)</u> | <u>Controls</u><br><u>(n=411)</u> |
| --- | --- | --- | --- |
| **Age** |  |  |  |
| 15-17 years | 81 (19.7%) | 25(6.1%) | 56(13.6%) |
| 18- 20 years | 68 (16.7%) | 33(8.0%) | 35(8.5%) |
| 21-24 years | 57 (13.9 %) | 20(4.9%) | 37(9.0%) |
| 25-30 years | 100 (24.3%) | 31(7.5%) | 69(16.8%) |
| 31-35 years | 105 (25.5%) | 28(6.8%) | 77(18.7%) |
| **Sex** |  |  |  |
| Male | 119 (29%) | 45(10.9%) | 74(18.0%) |
| Female | 292 (71%) | 92(22.4%) | 200(48.7%) |
| **Marital status** |  |  |  |
| Single | 157 (38.2%) | 54(13.1%) | 103(25.1%) |
| Married | 185 (45%) | 55(13.4%) | 130(31.6%) |
| Widowed | 15 (3.6%) | 8(1.9%) | 7(1.7%) |
| Separated/divorced | 54 (13.1%) | 20(4.9%) | 34(8.3%) |
| **Religion** |  |  |  |
| Christian | 369 (90%) | 126(30.7%) | 243(59.1%) |
| Moslem | 27 (6.6%) | 9(2.2%) | 18(4.4%) |
| Traditionalist | 14 (3.4%) | 2(0.2) | 12(2.9%) |

| **Education level** |  |  |  |
| --- | --- | --- | --- |
| None | 19 (4.6%) | 7(1.7%) | 12(2,9%) |
| Primary | 244 (59.4%) | 82(20.0% | 162(39.4%) |
| Secondary | 148 (36%) | 48(11.7%) | 100(24.3%) |

| <u>Variable</u> | <u>Frequency/Percentage</u> | <u>Cases</u><br><u>(n=411)</u> | <u>Controls</u><br><u>(n=411)</u> |
| --- | --- | --- | --- |
| **occupation** |  |  |  |
| House wife | 67 (16.3%) | 19(4.6%) | 48(11.7%) |
| Farmer | 68 (16.5%) | 18(4.4%) | 50(12.2%) |
| Skilled work | 39 (9.5%) | 14(3.4%) | 25(6.1%) |
| Small scale business | 62 (15.1%) | 22(5.4%) | 40(9.7%) |
| Student | 101 (24.6%) | 34(8.3%) | 67(16.3%) |
| Unemployed | 50 (12.2%) | 17(4.1%) | 33(8.0%) |
| Employed | 6 (1.5%) | 0(0%) | 6(1.5%) |
| Other | 17 (4.1%) | 12(2.9%) | 5(1.2%) |

| **Means of transport to the hospital** |  |  |  |
| --- | --- | --- | --- |
| By foot | 298 (72.5%) | 99(24.1%) | 199(48.4%) |
| By bicycle | 35 (8.5%) | 15(3.6%) | 20(4.9%) |
| By Car | 78 (19%) | 23(5.6%) | 55(13.4%) |

| **Time taken to get to the hospital** |  |  |  |
| --- | --- | --- | --- |
| Less than 30 minutes | 178 (43.3%) | 55(13.4%) | 123(29.9%) |
| More than 30 minutes but <1 hour | 138 (33.6%) | 38(9.2%) | 100(24.3%) |
| More than 1 hour | 86 (20.9%) | 44(10.7%) | 44(10.7%) |
| Unable to estimate | 1 (0.2%) | 0(0.0%) | 1(0.2%) |

### Relationship between ART defaulting and social economic factors

In table 3.0 findings suggest that a significant relationship was seen between ART defaulting and the following variables: participant’s age (X^2^=0.046, Cramer’s V: 0.2, OR: 0.663, 95% CL: 0.439-1.002); occupation (X^2^= 0.048, Cramer’s V: 0.1, OR: 0.658, 95% CL: 0.434-0.997) and how long the clients took to reach the clinic for ART services (X^2^= 0.002, Cramer’s V: 0.2, OR: 2.454, 95%CL: 1.511-3.985). Regarding specific reasons for missing hospital appointment, a significant relationship was seen between ART defaulting and migration to RSA (X^2^= 0.001, Cramer’s V: 0.270, OR: 35.661, 95% CL: 4.675-272.049).

**Table 3.0:**
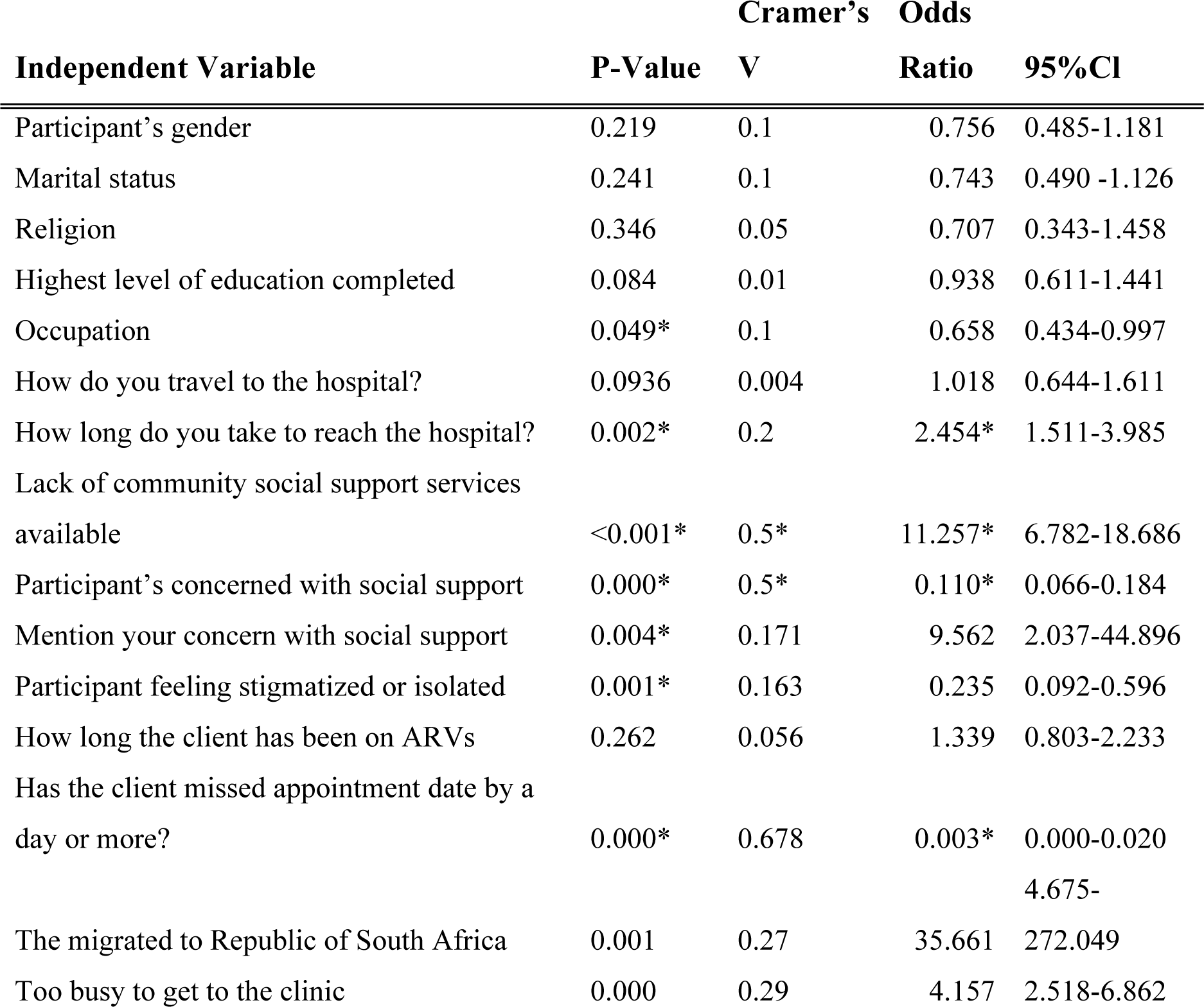
The relationship of community social support factors with ART defaulting.

### Major Predictors of ART Defaulting

From the table 4.0 above, the model shows a significant relationship (p value <0.05 on 95% level of significance. It can be suggested that the following community factors are the major predictors of ART defaulting in Mzimba District as depicted by this study are: long distances to the clinic (AOR=2.195, 95% CL: 1.186-4.061), lacking social support (AOR= 7.488, 95% CL: 3.486-14.570), and migration to RSA (AOR= 3.297, 95% CL: 1.728-6.292). This means that the youth are more likely to default ART if they travel long distances to the clinic, lacked social supportor migrated to RSA.

**Table 4.0.** Binary Logistic Regression Results.

| RISK FACTOR | UNADJUSTED |  | ADJUSTED |  |
| --- | --- | --- | --- | --- |
|  | OR (95% CI) | P-value | OR (95% CI) | P-value |
| Occupation | 0.658 (0.434-0.997) | 0.049 | 0.662 (0.389-1.127) | 0.128 |
| Long distance to the clinic | 2.454 (1.511-3.985) | 0.002 | 2.195 (1.186-4.061) | 0.012* |
| Lack of community social support | 11.257 (6.782-18.686) | <0.001 | 7.488 (3.486-14.570) | <0.001* |
| Stigma | 0.235 (0.092-0.596) | 0.001 | 1.005 (0.222-4.557) | 0.995 |
| Migration to RSA | 35.661 (4.675-272.049) | 0.001 | 3.297 (1.728-6.292) | <0.001* |
| Participant concerned with social support | 0.110 (0.066-0.184 ) | <0.001 | 6.856 (0.686-68.481) | 0.101 |

## Discussion

We conducted a case control study to determine factors associated with ART defaulting in Mzimba South. We found that ART defaulting is associated with a number of community social factors. These include: long distance to the clinic, occupation, lack of social support, stigma, migrationto RSA and participant concern with social support.

### ART Defaulting and Distance to Clinic

A statistically significant relationship between ART defaulting and long distance to the clinic was seen, (p: 0.002, OR: 2.454, 95% CL: 1.511-3.985). These findings imply that long distance to the clinic may influence ART defaulting. Similar observations were noted in earlier studies conducted in Malawi ^19,20^where ART defaulting was attributed to long distance to ART clinics. A possible explanation for these similarities might be due to common geographical barriers in Malawi such as poor roads and long distance to access health care services^21^. Although there is congruency in findings between the present study and the previous authors, it appears both studies considered the age group between 31-35 years as belonging to either youths or adults. This might have affected the outcome of both studies. Nonetheless, the findings are similar to another study in Nigeria which indicated that travel distances that exceed 2 hours to the health facility may influence the client to default ART^22^. These findings are in contrary to those of Chamberlain ^23^ who suggested that distance was not a barrier to retaining clients on ART program in Malawi. Reasons for these differences were not made clear in their study remain unclear. However, a related study conducted in Ethiopia noted that clients opted to attend distant ART clinics in order to have adequate privacy and avoid being recognised by community members from their society^24^.

In this study, clients were reported to behave like this to avoid stigma from people who knew them in the clinics near to them. There may be need to conduct more studies that explore determinants of ART defaulting in both nearby and distant ART clinics in Malawi. Nonetheless, the researchers in this present study suggest that long distance to ART clinic may influence ART defaulting among Malawian youths who cannot afford to pay for transport costs to the health facility on their appointment dates.

### ART Defaulting and Community Social Support

Lack of social support was observed to be significantly associated with ART defaulting among the youth (p-value: <0.001, OR: 11.257, 95% CL: 6.782-18.686). These results suggest that the odds of defaulting ART is 11.257 times higher among the youth that lack social support compared to those that have the needed support. The results are in accord with a study conducted in Malawi that assessed factors associated with adherence and retention in care of HIV positive clients^25^. The nature of ART programs desires daily taking of drugs, persevering to side effects and social stigma associated with the diagnosis^15^. Availability of social support such as constant reminders to the youth about scheduled visits to the clinic and encouragement, may enable clients to endure side effects and stigma ^26^. Lack of social support among the youth taking ARVs may leave them frustrated and remain with no drive to continue with treatment. In another study, social support inform of encouragement, food, money and transport improved medication adherence and quality of life among HIV-positive clients taking ART in many settings in Uganda ^27^. On the other hand, this social support may only be effective when the client trusts the community offering the support. The community care givers need to demonstrate confidentiality, love and concern for the youth. The client may feel comfortable with such community care providers. If the youth perceive their social support system as lacking confidentiality and trustworthiness in matters concerning their HIV status and care, the client may still default treatment ^7^.

### ART Defaulting and Migration to RSA

Results from this study indicate that there is a significant association between travelling to the republic of South Africa (RSA) and ART defaulting (p-value: 0.001, Cramer’s V: 0.270, OR: 35.661, 95% CL: 4.675-272.049). These findings suggest that the odds of defaulting ART treatment was 35.661 times higher among the HIV positive youth who travelled to South Africa to work, compared to those who did not. This finding broadly supports the work of other studies in this area linking a higher ART defaulting risk among migrants compared to non-migrants ^28^. This result may be explained by the fact that the youths could be defaulting ART because of defragmented ART service delivery across borders. For instance, a client registered in Malawi on ART program is free to collect drugs in his country ^29^. If this youth travels to RSA, it is alleged that foreign policy restrictions make ART drug access difficult ^28^. Similarly a South African newspaper recently revealed that most migrants fail to access ARVs because they lack national identity (ID) documents which are a requirement in public health institutions ^30^. ID requirement deprives the youth from timely accessing essential drugs like ARVs. Further, some of these youths might have expired IDs or might have gone to RSA without travel documents^31, 32^. On the other hand, it has been observed that these youths may avoid defaulting ART if their support system has been effective in collecting drugs for them. For instance, parents or guardians who consistently collect drugs on behalf of the youth from the clinic may lessen incidents of ART defaulting. Anecdotal evidence in Mzimba district has revealed that the ART is sent through transporters who frequently travel between Malawi and South Africa. In this practice, the clinic could dispense ART drugs for as much as 12 months to the guardian ^33^. ART records are then updated to show that the client is still on the program. Although this practice has been appraised in prevention of defaulting, it has some loop holes. ART drugs may not get to the intended client despite the relatives collecting from the clinic. This finding differs from other researchers^34^who posited that international migration did not have a statistical significance with defaulting and mortality among HIV clients in Brazil, Paraguay and Argentina. The differences would be due to supportive policies regarding access to ARVs towards migrants. We recommend on the need for a cross-border collaboration in managing clients on ART between countries to ensure continuity of care.

### What is the Key Finding From this Study?

ART defaulting among the youth living with HIV in Mzimba, Malawi, is influenced by several community factors. These are lack of community social support, long distance to clinic and migration to South Africa in search for greener pasture.

### Strengths and limitations

The sample size was large enough to increase representation of study participants to the population under study. The use of two-stage cluster random sampling ensured diverse representation of participants from all health facilities in Mzimba South. The findings of this study were collected in Mzimba South only, hence generalization of these findings to the whole district may be limited and results from this study should be utilised with caution. Nonetheless, the internal and external validity of the study was not adversely affected.

## Conclusions

ART defaulting contributes to development of resistant strains of HIV. The study has found that HIV positive youth in Mzimba South default ART treatment due to long distances to clinic, lack of social support and migration to RSA in search for employment.

Based on the study findings, the researcher has recommended the strengthening of mental health during pre-service and in-service nursing education. The nursing curriculum should be reviewed to generate competencies in screening and management of psychological disorders among youths and all clients on ART. Early detection and management of psychological disorders will help to improve retention of the youths in ART programmes.

In addition, there is need to introduce Community ART Outreach Programmes to be included under comprehensive health service delivery. The institution of Family-centered care that ensures identified family members are trained on rendering social support to youths on ART in their families. This would contribute to reducing defaulting among the youth.

Further, policy review to provide for technology that enables free ARVs regardless of national ID requirements and cross-border collaboration in managing HIV youths on ART between countries would ensure continuity of care

The researcher also recommended that more research on this subject be conducted in other parts of the country to ascertain its existence and severity. Nonetheless, the findings in this research may still need to be considered when planning care for the youths living with HIV in Mzimba South.

## Data Availability

All relevant data are within the manuscript and its Supporting Information files.

## Acknowledgements

We appreciate all youths from Mzimba District and Embangweni Hospital, Euthini and Jenda health centers for voluntarily participating in this study. Many thanks should go to the authorities of Mzimba South DHO for permitting us to conduct the study in their health facilities. In addition, we appreciate the College of Medicine, the Malawi Medical Journal and the Blantyre Institute for Community Outreach (BICO) who organized and sponsored manuscript writing workshop. This helped us to perfect ourwriting skills. We thank the Malawi Ministry of Education through the Higher Education Research Grants for funds that partly supported this study.

## References

1. Kim MH, Mazenga AC, Yu X, Devandra A, Nguyen C, Ahmed S, et al. Factors associated with depression among adolescents living with HIV in Malawi. BMC Psychiatry [Internet]. 2015 Oct 26 [cited 2019 Dec 1];15. Available from: https://www.ncbi.nlm.nih.gov/pmc/articles/PMC4624356/

2. Brown JP, Ngwira B, Tafatatha T, Crampin AC, French N, Koole O. Determinants of time to antiretroviral treatment initiation and subsequent mortality on treatment in a cohort in rural northern Malawi. AIDS Research and Therapy. 2016 Jul 8;13(1):24.

3. Global Health. Global Health | Malawi | U.S. Agency for International Development [Internet]. 2021 [cited 2021 Aug 7]. Available from: https://www.usaid.gov/malawi/global-health

4. MacPherson P, Munthali C, Ferguson J, Armstrong A, Kranzer K, Ferrand RA, et al. Service delivery interventions to improve adolescents’ linkage, retention and adherence to antiretroviral therapy and HIV care. Tropical medicine & international health. 2015;20(8):1015–32.

5. Mugglin C, Haas AD, Oosterhout JJ van, Msukwa M, Tenthani L, Estill J, et al. Long-term retention on antiretroviral therapy among infants, children, adolescents and adults in Malawi: A cohort study. PLOS ONE. 2019 Nov 14;14(11):e0224837.

6. Ahmed A, Dujaili JA, Jabeen M, Umair MM, Chuah LH, Hashmi FK, et al. Barriers and Enablers for Adherence to Antiretroviral Therapy Among People Living With HIV/AIDS in the Era of COVID-19: A Qualitative Study From Pakistan. Frontiers in pharmacology. 2021;3968.

7. Kim MH, Zhou A, Mazenga A, Ahmed S, Markham C, Zomba G, et al. Why Did I Stop? Barriers and facilitators to uptake and adherence to ART in option B+ HIV care in Lilongwe, Malawi. PLOS ONE. 2016 Feb 22;11(2):e0149527.

8. McGuire M, Munyenyembe T, Szumilin E, Heinzelmann A, Paih ML, Bouithy N, et al. Vital status of pre-ART and ART patients defaulting from care in rural Malawi. Tropical Medicine & International Health. 2010 Apr 29;15:55–62.

9. Ministry of Health. The Malawi guidelines for clinical management of HIV in children and adults. Ministry of Health; 2016.

10. Xing H, Ruan Y, Li J, Shang H, Zhong P, Wang X, et al. HIV drug resistance and its impact on antiretroviral therapy in Chinese HIV-infected patients. PloS one. 2013;8(2):e54917.

11. Lay PRD, Benzaken A, Karim QA, Aliyu S, Amole C, Ayala G, et al. Ending AIDS as a public health threat by 2030: Time to reset targets for 2025. PLOS Medicine. 2021 Jun 8;18(6):e1003649.

12. Adejumo OA, Malee KM, Ryscavage P, Hunter SJ, Taiwo BO. Contemporary issues on the epidemiology and antiretroviral adherence of HIV-infected adolescents in sub-Saharan Africa: a narrative review. Journal of the International AIDS Society. 2015;18(1):20049.

13. Zhou A. The uncertainty of treatment: Women’s use of HIV treatment as prevention in Malawi. Social Science & Medicine. 2016 Jun 1;158:52–60.

14. Chirambo L, Valeta M, Banda Kamanga TM, Nyondo-Mipando AL. Factors influencing adherence to antiretroviral treatment among adults accessing care from private health facilities in Malawi. BMC Public Health. 2019 Oct 28;19(1):1382.

15. Renju J, Moshabela M, McLean E, Ddaaki W, Skovdal M, Odongo F, et al. ‘Side effects’ are ‘central effects’ that challenge retention in HIV treatment programmes in six sub-Saharan African countries: a multicountry qualitative study. Sex Transm Infect [Internet]. 2017 Jul 1 [cited 2022 Jan 5];93(Suppl 3). Available from: https://sti.bmj.com/content/93/Suppl_3/e052971

16. Cataldo F, Chiwaula L, Nkhata M, van Lettow M, Kasende F, Rosenberg NE, et al. Exploring the Experiences of Women and Health Care Workers in the Context of PMTCT Option B Plus in Malawi. J Acquir Immune Defic Syndr. 2017 Apr 15;74(5):517–22.

17. Elwell K. Facilitators and barriers to treatment adherence within PMTCT programs in Malawi. AIDS Care. 2016 Aug 2;28(8):971–5.

18. Republic of South Africa. New requirement at clinics in South Africa causes panic amongst HIV+ foreign nationals [Internet]. Regional Interagency Task Team on Children Affected by AIDS. 2017 [cited 2020 Feb 7]. Available from: http://www.riatt-esa.org/news-1/2018/2/12/new-requirement-at-clinics-in-south-africa-causes-panic-amongst-hiv-foreign-nationals

19. Bilinski A, Birru E, Peckarsky M, Herce M, Kalanga N, Neumann C, et al. Distance to care, enrollment and loss to follow-up of HIV patients during decentralization of antiretroviral therapy in Neno District, Malawi: A retrospective cohort study. PLOS ONE. 2017 Oct 3;12(10):e0185699.

20. Chirambo L, Valeta M, Kamanga TMB, Nyondo-Mipando AL. Factors influencing adherence to antiretroviral treatment among adults accessing care from private health facilities in Malawi. BMC public health. 2019;19(1):1–11.

21. Munthali AC, Swartz L, Mannan H, MacLachlan M, Chilimampunga C, Makupe C. “This one will delay us”: barriers to accessing health care services among persons with disabilities in Malawi. Disability and rehabilitation. 2019;41(6):683–90.

22. Chime OH. Rates and predictors of adherence and retention for antiretroviral therapy among HIV-positive adults in Enugu, Nigeria. Malawi Medical Journal. 2019;31(3):202–11.

23. Chamberlin S, Mphande M, Phiri K, Kalande P, Dovel K. How HIV clients find their way back to the ART clinic: A Qualitative study of disengagement and re-engagement with HIV care in Malawi. AIDS Behav [Internet]. 2021 Aug 17 [cited 2021 Oct 4]; Available from: https://doi.org/10.1007/s10461-021-03427-1

24. Lifson AR, Demissie W, Tadesse A, Ketema K, May R, Yakob B, et al. Barriers to retention in care as perceived by persons living with HIV in rural Ethiopia: focus group results and recommended strategies. Journal of the International Association of Providers of AIDS Care (JIAPAC). 2013;12(1):32–8.

25. Gugsa S, Potter K, Tweya H, Phiri S, Sande O, Sikwese P, et al. Exploring factors associated with ART adherence and retention in care under Option B+ strategy in Malawi: A qualitative study. PLOS ONE. 2017 Jun 21;12(6):e0179838.

26. Phiri S, Tweya H, van Lettow M, Rosenberg NE, Trapence C, Kapito-Tembo A, et al. Impact of facility-and community-based peer support models on maternal uptake and retention in Malawi’s option B+ HIV prevention of mother-to-child transmission program: a 3-arm cluster randomized controlled trial (PURE Malawi). JAIDS Journal of Acquired Immune Deficiency Syndromes. 2017;75:S140–8.

27. Atukunda EC, Musiimenta A, Musinguzi N, Wyatt MA, Ashaba J, Ware NC, et al. Understanding Patterns of Social Support and Their Relationship to an ART Adherence Intervention Among Adults in Rural Southwestern Uganda. AIDS and Behavior. 2017;21(2):428.

28. Luke International. Luke international - Cross-border challenges [Internet]. Luke international. 2021 [cited 2021 Aug 9]. Available from: https://lukeinternational.no/cross-border-challenges/

29. Dickerson S, Baranov V, Bor J, Barofsky J. Treatment as insurance: HIV antiretroviral therapy offers financial risk protection in Malawi. Health Policy and Planning. 2020 Jul 1;35(6):676–83.

30. Chinele J. Malawi ARVs traded on Jozi’s streets [Internet]. The Mail & Guardian. 2015 [cited 2021 Aug 9]. Available from: https://mg.co.za/article/2015-12-03-malawi-arvs-traded-on-jozis-streets/

31. Mtonga L. Malawi: How Malawians Travel to South Africa Without Documents. Nyasa Times [Internet]. 2019 Dec 18 [cited 2022 Apr 21]; Available from: https://allafrica.com/stories/201912180678.html

32. Agencia de Informacao de Mocambique. Mozambique: 24 Illegal Malawian Migrants Detained. Agencia de Informacao de Mocambique [Internet]. 2021 May 7 [cited 2022 Apr 21]; Available from: https://allafrica.com/stories/202105070683.html

33. Ministry of Health. Malawi Clinical HIV guidelines 2018. Ministry of Health; 2018.

34. Zaslavsky R, Goulart BNG de, Ziegelmann PK. Cross-border healthcare and prognosis of HIV infection in the triple border Brazil-Paraguay-Argentina. Cad Saúde Pública [Internet]. 2019 Sep 9 [cited 2021 Aug 9];35. Available from: http://www.scielo.br/j/csp/a/9XHKb9vSzDYnYXybrMqrMPn/?lang=en

